# Impact of SARS-CoV-2 on the microbiota of pregnant women and their infants

**DOI:** 10.1101/2022.11.01.22281810

**Authors:** Heidi K. Leftwich, Daniela Vargas-Robles, Mayra Rojas-Correa, Yan Rou Yap, Shakti Bhattarai, Doyle V. Ward, Gavin Fujimori, Catherine S. Forconi, Tracy Yeboah, Acara Carter, Alyssa Kastrinakis, Alison M. Asirwatham, Vanni Bucci, Ann M. Moormann, Ana Maldonado-Contreras

## Abstract

The microbiome inherited at birth exerts marked effects on immune programming with long-term health consequences. Here, we demonstrated that the gut, vaginal, and oral microbial diversity of pregnant women with SARS-CoV-2 infection is reduced, and women with early infections exhibit a different vaginal microbiota composition compared to healthy controls at the time of delivery. Accordingly, infants born to pregnant women with early SARS-CoV-2 infection exhibit a unique oral microbiota dominated by *Streptococcus* species. Together, we demonstrated that SARS-CoV-2 infections during pregnancy, particularly early infections, are associated with lasting changes in the microbiome of pregnant women compromising the initial microbial seed of their infant. Our results highlight the importance of further exploring the impact of SARS-CoV-2 on the infant’s microbiome-dependent immune programming.

**One Sentence Summary:** Pregnant patients with SARS-CoV-2 infection early in pregnancy and with active infection exhibit an altered vaginal and oral microbiota that is passed on to infants.

## INTRODUCTION

Every infant “inherits” their first gut bacteria from their mother (*1-6*). The inherited microbiome exerts marked effects on immune programming with long-term health consequences, including susceptibility to infections or chronic inflammatory diseases and reduced vaccine efficacy (*7-14*). Particularly, epidemiological and mechanistic studies in animal models have established that microbial dysbiosis in early life influences disease pathogenesis via changes in immune system maturation (*12, 13, 15*). Many acute and chronic diseases have now been associated with changes in the oral and gut microbiomes, and there is evidence of the complex interplay between the immune system, systemic physiology, and the microbiome in various health conditions. Therefore, this “window of opportunity” at birth, either renders infants with a healthy immune system or alternatively establishes a divergent path leading to severe immune-mediated disease susceptibility (*16-24*). The impact of SARS-CoV-2 infections on pregnant women and their offspring microbiotas has not been fully explored.

Pregnant women are of particular interest as poor perinatal outcomes related to SARS-CoV-2 infection have been reported (*25-28*). Pregnant women can manifest severe symptoms of SARS-CoV-2 infection (*27, 29*). Particularly, these patients have increased risks of mechanical ventilation, extracorporeal membrane oxygenation (ECMO), intensive care unit admission, hypertensive disorders of pregnancy, thrombotic disease, and death (*27, 29*). There are also studies showing the effects of maternal SARS-CoV-2 infection on the fetus such as the increased risk of preterm birth, intrauterine growth restriction, and higher cesarean delivery rate (*27, 29-31*).

Most studies exploring the impact of SARS-CoV-2 infection on the microbiota have focused on the lung (*32*), gut (*33-35*), and nasopharyngeal mucosa (*36*) in non-pregnant adults. Two studies to our knowledge, have characterized the gut and nasopharyngeal microbiotas at the time of delivery and post-partum colostrum of pregnant women with SARS-CoV-2 infection (*37, 38*). The first study conducted in Spain, reports an increased abundance of Bacteroidales in the nasopharyngeal swabs of pregnant women with active SARS-CoV-2 infection compared to healthy controls. The second study authors observed differences in the gut microbiota of stool positive pregnant women and their infants compared to the pregnant women with no evidence of SARS-CoV-2 viral particles in stool. Most of the pregnant women included in the study were negative for SARS-CoV-2 on nasopharyngeal swabs and asymptomatic at time of sample collection, suggesting past SARS-CoV-2 infection.

Besides gut microbiota, the vaginal and oral microbiotas of pregnant women represent the initial seed of the infants’ gut microbiota if born vaginally (*1, 3*). There is a paucity of research in understanding the impact of SARS-CoV-2 infection at different stages of pregnancy on the initial seed of the infant’s microbiota (i.e., gut, vaginal, and oral microbiotas of pregnant women). Hence, the objective of this study is to determine whether infection by SARS-CoV-2 during pregnancy, either at early or late stages of pregnancy, or an active infection at delivery results in gut, vaginal, and oral microbiota changes that are passed onto the offspring.

## RESULTS

### Participant description

We recruited 62 pregnant women with SARS-CoV-2 infection and 38 healthy pregnant controls (**Table 1**). One of the participants in the healthy control group showed higher nucleocapsid protein (NP) IgG OD than the cutoff established by us (>0.57 OD) (*39*), thus was excluded from the analyses. Therefore, a total of 99 pregnant women, 61 with SARS-CoV-2 infection, and 38 healthy pregnant controls were included in the analyses. Participants had an average age of 32 years, and a pre-pregnancy body mass index (BMI) of 30.3, consistent with a diagnosis of obesity. In fact, 41.0% of participants were obese by pre-pregnancy BMI, while 16.2% exhibited a normal BMI.

**Table 1.** Demographic and clinical descriptions of the pregnant participants included in the study. SARS-CoV-2 positive participants and SARS-CoV-2 negative (healthy controls) were recruited at the University of Massachusetts Memorial Hospital from April 27 to June 10, 2020. Additional, SARS-CoV-2 negative (healthy controls) were recruited nationwide prior to the COVID-19 pandemic.

| Demographics and clinical variables | SARS-CoV-2 positive pregnant women (N=62) | Healthy controls pregnant women (N=37) | TOTAL (N=99) | P value<sup>&</sup> |
| --- | --- | --- | --- | --- |
| **Age** |  |  |  | 0.001 |
| Mean (SD) | 31.0 (6.24) | 33.7 (4.79) | 32.0 (5.86) |  |
| **Pre-pregnancy body mass index** |  |  |  |  |
| Mean (SD) | 31.2 (6.93) | 28.7 (6.78) | 30.3 (6.95) |  |
| Missing | 0 (0%) | 1 (2.7%) | 1 (1.0%) |  |
| **Race** |  |  |  | 0.001 |
| Non-Hispanic White | 29 (46.8%) | 28 (75.7%) | 57 (57.6%) |  |
| Hispanic or Latino | 24 (38.7%) | 2 (5.4%) | 26 (26.3%) |  |
| Non-Hispanic Black | 7 (11.3%) | 4 (10.8%) | 11 (11.1%) |  |
| Non-Hispanic Asian | 2 (3.2%) | 3 (8.1%) | 5 (5.1%) |  |
| **SARS-CoV-2 comorbidities** |  |  |  |  |
| Type 2 diabetes |  |  |  | 0.863 |
| Yes | 4 (6.5%) | 2 (5.4%) | 6 (6.1%) |  |
| Missing | 0 (0%) | 10 (27.0%) | 10 (10.1%) |  |
| Cardiovascular disease |  |  |  | 0.89 |
| Yes | 12 (19.4%) | 5 (13.5%) | 17 (17.2%) |  |
| Missing | 0 (0%) | 10 (27.0%) | 10 (10.1%) |  |
| **Pregnancy outcomes** |  |  |  |  |
| Preeclampsia |  |  |  | 0.557 |
| Yes | 9 (14.5%) | 3 (8.1%) | 12 (12.1%) |  |
| Missing | 1 (1.6%) | 0 (0%) | 1 (1.0%) |  |
| Preterm 37 weeks |  |  |  | 0.305 |
| Yes | 14 (22.6%) | 5 (13.5%) | 19 (19.2%) |  |
| Gestational diabetes |  |  |  | 0.153 |
| Yes | 13 (21.0%) | 3 (8.1%) | 16 (16.2%) |  |
| Antibiotic during delivery |  |  |  | 0.157 |
| Yes | 24 (38.7%) | 12 (32.4%) | 36 (36.4%) |  |
| Missing | 0 (0%) | 10 (27.0%) | 10 (10.1%) |  |
| Antibiotic before delivery |  |  |  | 0.644 |
| Yes | 5 (8.1%) | 0 (0%) | 5 (5.1%) |  |
| **Vaccinated against SARS-CoV-2** |  |  |  |  |
| Yes | 1 (1.6%) | 9 (24.3%) | 10 (10.1%) | 0.005 |

& Fisher's exact test for categorical variables and Kruskal-Wallis test for continuous variables

Demographic and health comparisons revealed that age and the prevalence of non-Hispanic white women were significantly higher in the healthy control group (33.7 years old and 75.7% respectively) than in the SARS-CoV-2 group (**Table 1** and **Table S1**, P<0.002). As most of our recruitment occurred before vaccines against SARS-CoV-2 became available, only a fraction of the participants (10.1%) were vaccinated at the time of sample collection. The COVID-19 vaccination rate was higher in the healthy control group compared to SARS-CoV-2 positive participants (**Table 1** and **Table S1**, P<0.005).

The infants recruited for the study were 52.0% female, with a mean of 3,280 grams of body weight, and most were delivered vaginally (68.8%). A total of 15.6% of infants were admitted to the newborn intensive care unit mostly due to pre-term birth (P<0.0001), and not associated with SARS-CoV-2 infection during pregnancy (P=0.278) or with active infections at delivery (P=0.436). There were no differences in demographic, or anthropometrics between infants born to SARS-CoV-2 positive or healthy control women (**Table 2**, P>0.050). The rate of cesarean was significantly higher (60.0%) for active SARS-CoV-2 at delivery compared to other groups (**Table S2**, P=0.029).

**Table 2.**
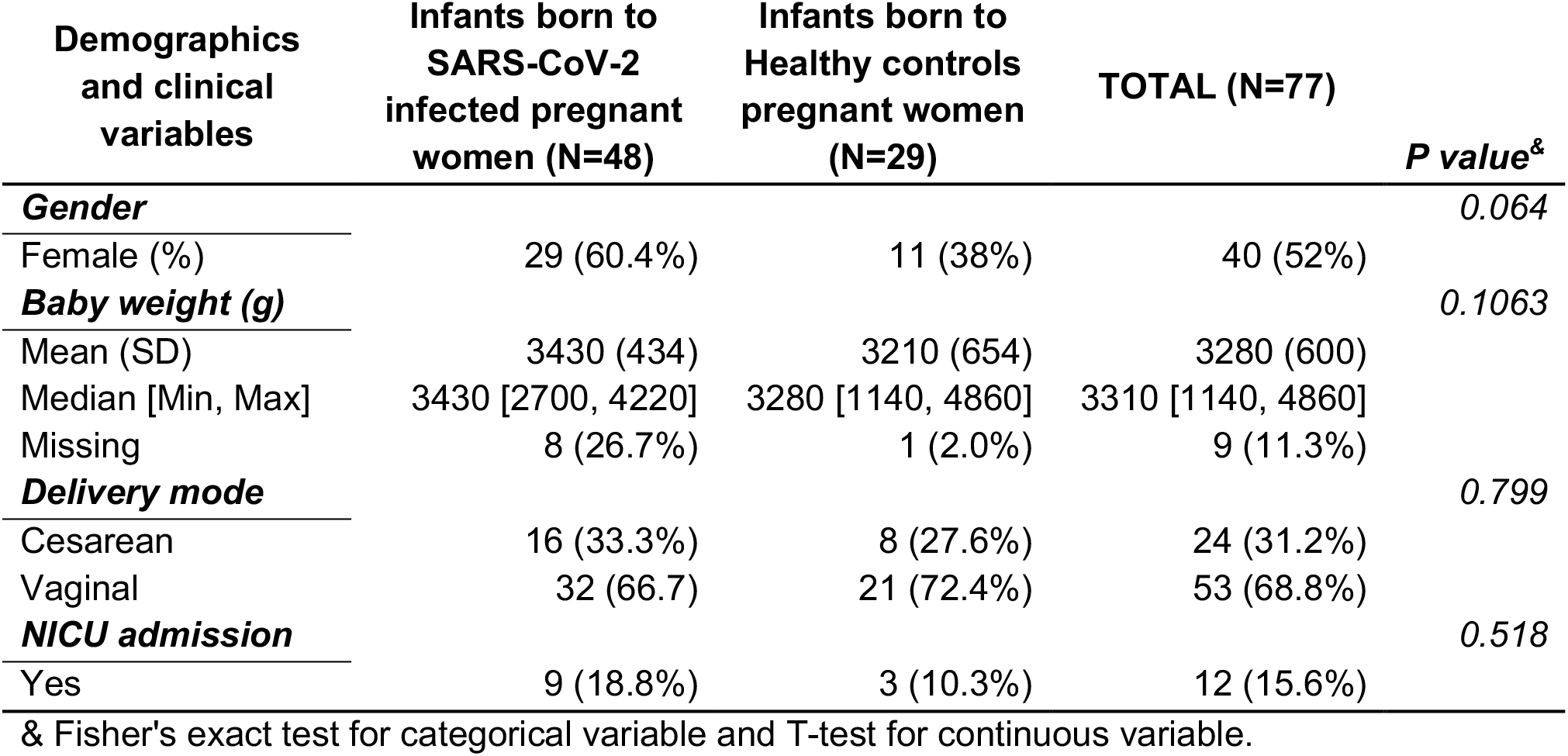
Demographic and clinical descriptions of infants included in the study. Infants born to SARS-CoV-2 infected pregnant participants and healthy controls were recruited at the University of Massachusetts Memorial Health from April 27 to June 10, 2020. Additional infants born to healthy controls were recruited nationwide prior to the COVID-19 pandemic.

### SARS-CoV-2 infection is associated with changes on the microbiota composition of pregnant women

We evaluated the microbial diversity of pregnant women with SARS-CoV-2 infection during pregnancy and compared it to healthy controls; we also compared microbial diversity by the time of SARS-CoV-2 positive diagnosis, namely: early (1^st^ and 2^nd^ trimester of pregnancy), late (3^rd^ trimester of pregnancy), and active (at the time of delivery). Regardless of the timing of the SARS-CoV-2 positive diagnosis, all the samples were collected from pregnant women at delivery admission, prior to delivery. Alpha and beta diversity assessments, and Random Forest Classification (RFC) were performed including potential cofounder variables such as race, antibiotic use, mother’s age, pre-pregnancy BMI, and gestational diabetes.

#### Gut microbiota of pregnant women

Out of the 99 pregnant women included in the analyses, we obtained stool samples from 46 pregnant women with positive SARS-CoV-2 diagnoses and 23 from healthy pregnant controls (**Table S3**). Alpha diversity analyses showed that being diagnosed with SARS-CoV-2 during pregnancy was associated with lower gut microbial richness within samples compared to healthy controls (**Figure 1, Table S4**, Shannon index, Linear regression, P=0.015). Gut microbial richness did not differ by the timing of SARS-CoV-2 infection (**Table S5**, Chao1 estimator, Linear regression, P=0.015, Pairwise Estimated Marginal Means, Padj>0.050). There were no microbial compositional or inter-person variability (beta diversity) by SARS-CoV-2 infection (**Table S6**, PERMANOVA and Betadisper test, P>0.079).

**Figure 1.**
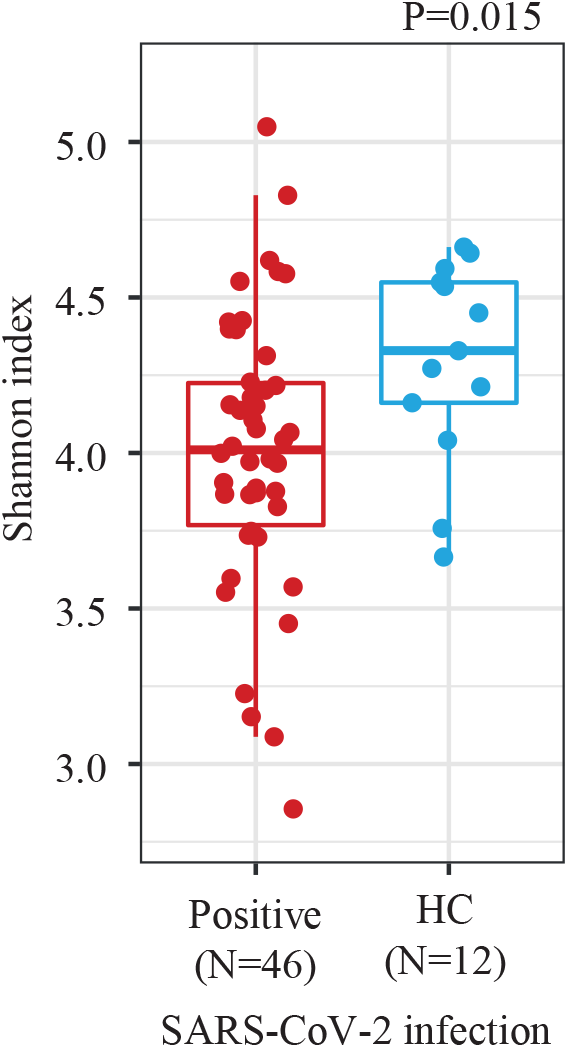
Gut microbial diversity of pregnant women differs by SARS-CoV-2 infection. Alpha diversity is shown using Shannon index comparing pregnant women with SARS-CoV-2 infection during pregnancy *vs*. pregnant women healthy controls (HC). The statistical test applied was a Linear regression.

We observed that gestational diabetes was significantly associated with higher gut microbial diversity (**Table S4 and S5**, Shannon, Linear regression, P<0.001), and the gut microbiota composition varied by pre-pregnant BMI in this cohort (Sørensen index, PERMANOVA, R^2^=2.6%, P=0.014).

We applied RFC to select the variables (taxa or demographic variables) that predict SARS-CoV-2 infection and then we used LIME algorithm to set abundance threshold values that best separate the two outcome groups. We found that SARS-CoV-2 infections were predicted (F1-score: 0.93) by a higher abundance of Selenomonadales (*Dialister*) and lower of Acidaminococcales (*Phascolarctobacterium faecium*), Eubacteriales (*Anaerostipes*), Bacteroidales (*Prevotella buccalis, Porphyromonas uenonis*, and *Bacteroides*) (see the taxa relative abundance threshold in **Figure S1A)**.

In sum, SARS-CoV-2 infection during pregnancy reduces the gut microbial diversity at delivery regardless of the timing of the diagnosis. Pregnant women infected with SARS-CoV-2 exhibit a decreased abundance of Acidaminococcales, Eubacteriales, Bacteroidales, and Selenomonadales compared to pregnant healthy controls.

### Vaginal microbiota of pregnant women

A total of 64 women were sampled for vaginal microbiota analyses. We obtained 43 samples from pregnant women with SARS-CoV-2 infection and 21 samples from healthy controls (**Table S3**). First, we classified the vaginal microbiota into Community State Types (CSTs) as previously done (*40*). Each CST is characterized by the dominance of a specific specie of *Lactobacillus* (i.e., *L. crispatus, L. gasseri, L. iners, or L. jensenii*) or the absence of *Lactobacillus*-dominance (*40*). Most of the pregnant women recruited for this study were classified on the *L. crispatus*-dominated profile (CST-I, 42.2%) followed by *L. iners*-dominated profile (CST-III, 37.5%). We did not find significant differences in CSTs distribution among SARS-CoV-2 positive pregnant women or by the time of diagnosis compared to healthy controls (**Table S7**, Fisher’s exact test, P>0.050).

We observed that pregnant women with SARS-CoV-2 infection exhibited lower vaginal microbial richness (**Figure 2A**, Chao1 estimator, Linear regression, P=0.005) compared to healthy controls. Moreover, pregnant women with early SARS-CoV-2 infection exhibited the lowest richness among all the pregnant women with SARS-CoV-2 infection when compared to healthy controls (**Figure 2B**, Pairwise Estimated Marginal Means, Padj=0.042).

**Figure 2.**
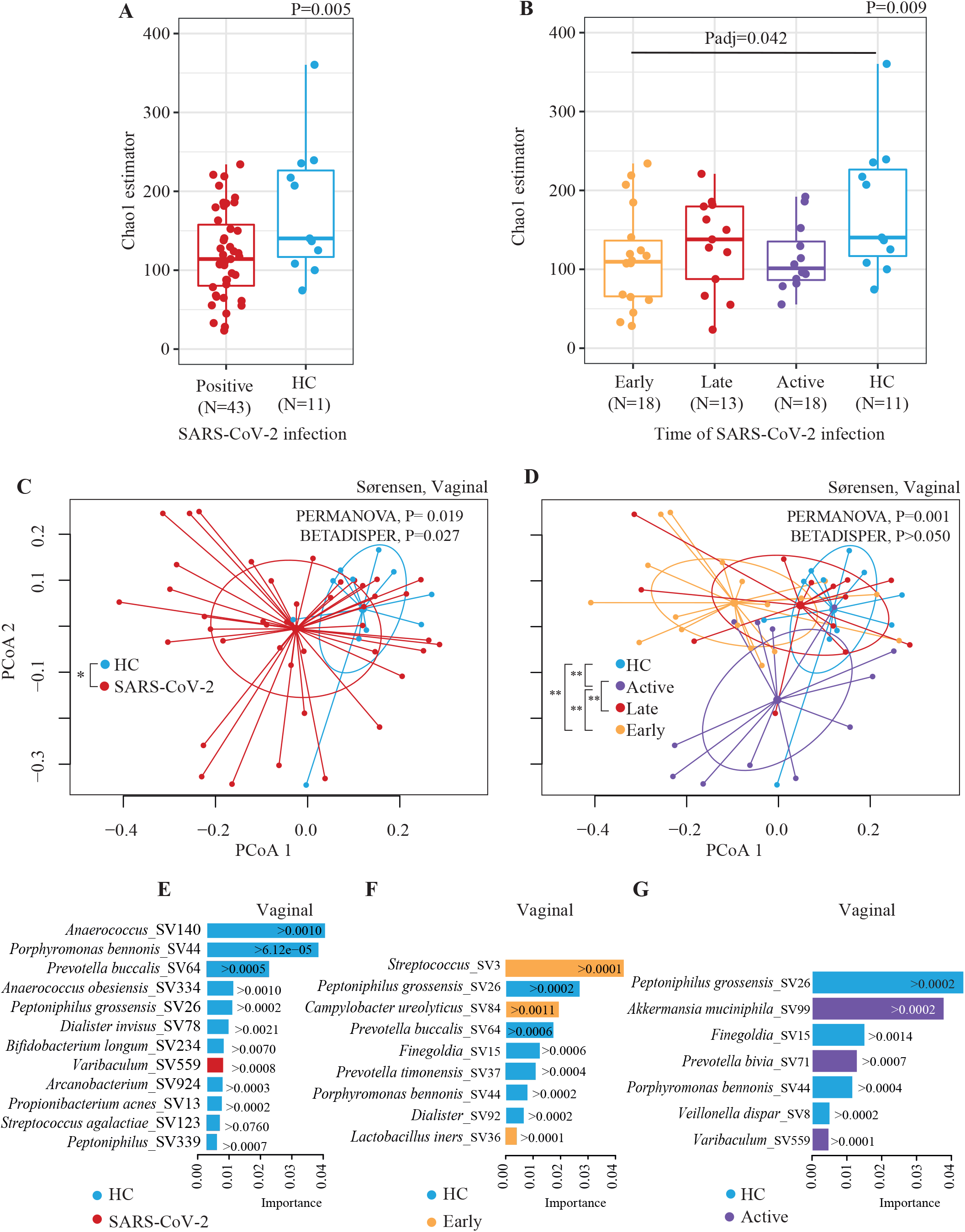
Vaginal microbiota of pregnant women differs by SARS-CoV-2 infection. (**A**) Alpha diversity is shown using Chao1 estimator comparing pregnant women with SARS-CoV-2 infection during pregnancy and pregnant women healthy controls (HC). (**B**) Alpha diversity is shown using Chao1 estimator comparing pregnant women with SARS-CoV-2 infection early, late, or active *vs*. HC. The statistical test applied was a Linear regression and pairwise comparison with Estimated Marginal Means. (**C** and **D**) Beta diversity analyses by groups: (**C**) Pregnant women with SARS-CoV-2 infection during pregnancy compared to HC; (**D**) pregnant women with SARS-CoV-2 infection early, late, or active compared to HC. Beta diversity comparisons were performed using PERMANOVA analysis with pairwise comparisons, and BETADISPER for dispersion analysis. For PERMANOVA and BETADISPER analyses we used Sørensen dissimilarities. P values were all adjusted by False Discovery Rate. (**E, F and G**) Bacterial taxa (at the amplicon sequence variant or SV) selected by the Random Forest Classification (RFC) and ranked according to their importance in the classification. RFC comparisons are shown in: (**E**) Pregnant women with SARS-CoV-2 infection during pregnancy *vs*. HC, (**F**) Pregnant women with early SARS-CoV-2 infection t *vs*. HC, (**G**) Pregnant women with active SARS-CoV-2 infection *vs*. HC. Bars’ colors indicate the comparison group (i.e., SARS-CoV-2 or HC); and each bar indicates the importance by which the increase on an SV predicts a particular comparison group. The selection of the variables for RFC was performed with Boruta algorithm. We also used the Local Interpretable Model-agnostic Explanation (LIME) to estimate a threshold of the abundance of the SV selected with Boruta that predicts a particular comparison group. *Padj<0.050,**Padj<0.010

We also observed that pregnant women with SARS-CoV-2 infection exhibited a microbiota composition distinct from healthy controls, particularly those with early or active infections, clustering the furthest from that group (**Figure 2C** and **D**, Sørensen index, PERMANOVA, R^2^=2.6%, and 8.6%, respectively P<0.050). In addition, we observed that participants with SARS-CoV-2 infection showed higher inter-person vaginal microbiota variability than healthy controls who overall exhibited a more similar microbiota composition among individuals (**Figure 2C** and **D, Table S6**, Sørensen index, BETADISPER analysis, adjusted for sample size differences, P=0.027). Of note, differences in microbiota composition were detected only when evaluated with the Sørensen index (unweighted measurement), but not with Bray-Curtis dissimilarity (weighted measurement), indicating that the compositional changes are mainly driven by rare microbial taxa.

As expected, the administration of antibiotics during delivery showed a significant effect on the vaginal microbiota diversity in this cohort (Sørensen index, PERMANOVA, R^2^=6.0%, P=0.030). However, antibiotic use was similar among infected and healthy control groups; thus, the effect of SARS-CoV-2 on the vaginal microbiota is relevant despite antibiotic use (**Table 1**, and **Table S1**).

Specifically, we found that SARS-CoV-2 infections were predicted (F1-score: 0.94) by a higher abundance of Actinomycetales (*Varibaculum*) and by a lower abundance of Clostridiales (*Anaerococcus sp*.), Bacteroidales (*P. bennonis*, and *P. buccalis*), Eubacteriales (*Peptoniphilus sp*.), Selenomonadales (*D. invisus*), Bifidobacteriales (*B. longum*), Actinomycetales (*Arcanobacterium*), Propionibacteriales (*P. acnes*), Lactobacillales (*S. agalactiae*) in the vaginal samples (see the taxa relative abundance threshold in **Figure 2E**).

Furthermore, we investigated whether the timing of SARS-CoV-2 infection impacted the vaginal microbiota composition. RFC and LIME algorithms determined that pregnant women with SARS-CoV-2 infection in early pregnancy were predicted (F1-score: 0.91) by a higher abundance of Lactobacillales (*Streptococcus* and *L. iners*), and Campylobacterales (*C. ureolyticus*) and by a lower abundance of different Eubacteriales (*P. grossensis, Finegoldia*, and *P. bennonis*), Bacteroidales (*P. buccalis* and *P. timonensis*), and Selenomonadales (*Dialister*) (**Figure 2F**). Pregnant women with active SARS-CoV-2 infection were predicted (F1-score: 0.83) by a higher abundance of Verrucomicrobiales (*A.muciniphila*), Bacteroidales (*P. bivia*) and Actinomycetales (*Varibaculum*), and by a lower abundance of Eubacteriales (*P. grossensis*), Clostridia (*Finegoldia*), Vellionellales (*V. dispar*) and Bacteroidales (*P. bennonis*) (**Figure 2G**). No taxon was predictive of pregnant women with late SARS-CoV-2 infection.

#### Oral microbiota of pregnant women

Out of the 79 pregnant women providing oral samples, 53 were positive for SARS-CoV-2 during pregnancy and were 26 healthy controls (**Table S3**). There were no alpha diversity differences between the groups (**Table S4, S5**, Shannon index or Chao1 estimator, Linear regression, P>0.050). However, the oral microbiota composition was better explained by race (PERMANOVA, R^2^=4.7%, P=0.023), followed by SARS-CoV-2 infection (**Figure 3A**. PERMANOVA, R^2^=4.5%, P=0.019). Particularly, pregnant women with active SARS-CoV-2 infection have significantly different oral microbiota compared to healthy controls (**Figure 3B**, Sørensen index, Pairwise PERMANOVA, Padj=0.042).

**Figure 3.**
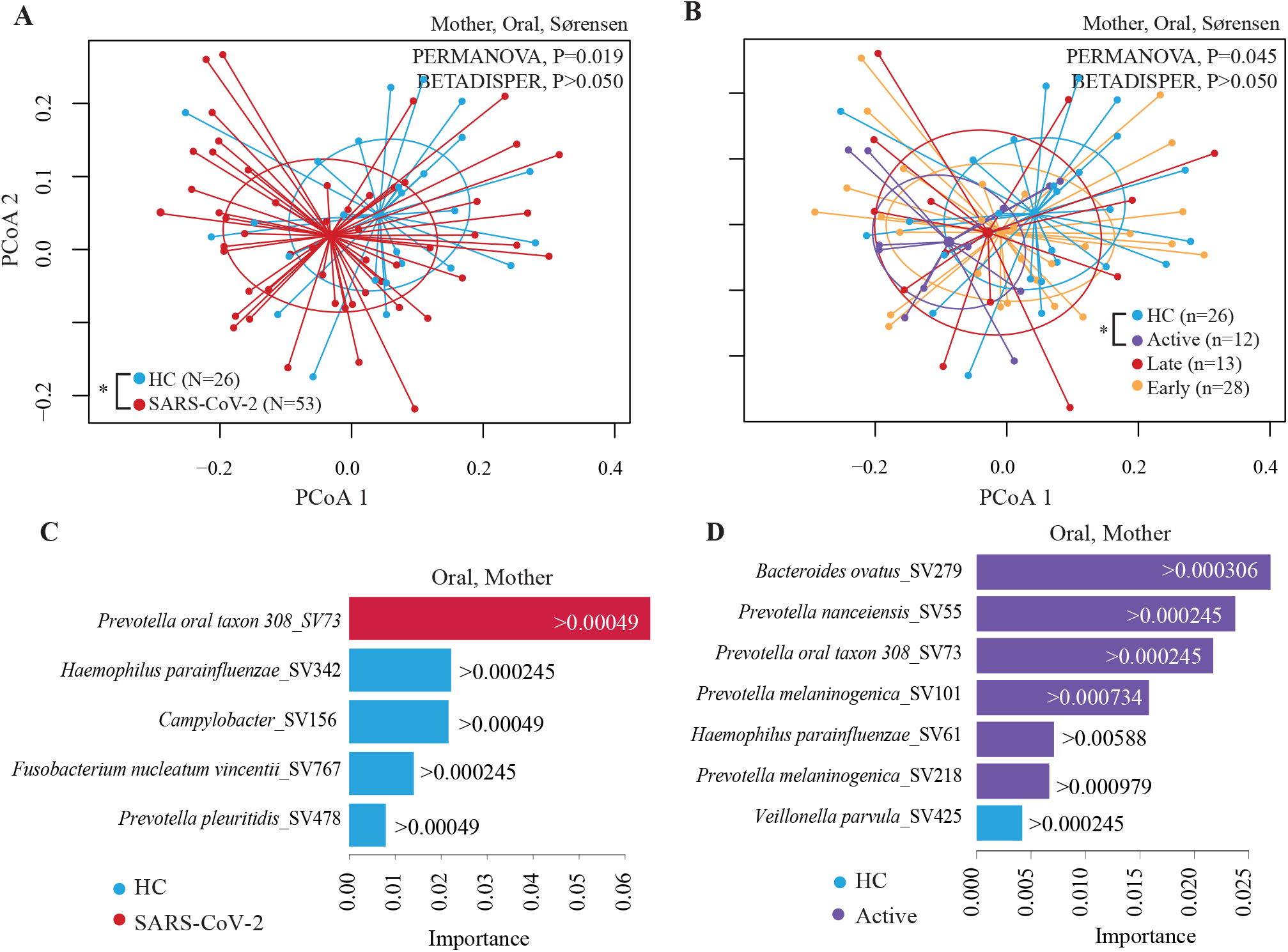
Oral microbiota of pregnant women differs by SARS-CoV-2 infection. Beta diversity analyses by groups: (**A**) Pregnant women with SARS-CoV-2 infection compared to HC; or (**B**) Pregnant women with early, late, or active SARS-CoV-2 infections compared to HC. Beta diversity comparisons were performed using PERMANOVA analysis with pairwise comparisons, and BETADISPER for dispersion analysis. For PERMANOVA and BETADISPER analyses we used Sørensen dissimilarities. P values were all adjusted by False Discovery Rate. (**C** and **D**) Bacterial taxa (at the amplicon sequence variant or SV) selected by the Random Forest Classification (RFC) and ranked according to their importance in the classification. RFC comparisons are shown in: (**C**) Pregnant women with SARS-CoV-2 infection compared to HC; (**D**) Pregnant women with active SARS-CoV-2 infection compared to HC. Bars’ colors indicate the comparison group (i.e., SARS-CoV-2 or HC); and each bar indicates the importance by which the increase on an SV predicts a particular comparison group. The selection of the variables for RFC was performed with Boruta algorithm. We also used the Local Interpretable Model-agnostic Explanation (LIME) to estimate a threshold of the abundance of the SV selected with Boruta that predicts a particular comparison group. *Padj<0.050, **Padj<0.010

RFC and LIME algorithms showed that the oral microbiota of pregnant women with SARS-CoV-2 infection were predicted (F1-score: 0.92) by a higher abundance of Bacteroidales (*Prevotella* oral taxon 308) and lower abundance of Pasteurellales (*H. parainfluenzae*), Campylobacterales (*Campylobacter*), Fusobacteriales (*F. nucleatum*), Bacteroidales (*P. pleuritidis*) (**Figure 3C**). Active SARS-CoV-2 infection were predicted (F1-score=0.87) by a higher abundance of Bacteroidales (*B. ovatus, P. nanceiensis*, oral taxon 308, *P. melaninogenica*) and Pasteurellales (*H. parainfluenzae*) and by a lower abundance of Clostridia (*Veillonella parvula*) (**Figure 3D**).

Altogether, the oral microbiota composition of pregnant women was remarkably different for pregnant women with SARS-CoV-2 infection compared to healthy controls.

### SARS-CoV-2 infection during pregnancy is associated with alterations in the microbiota of the offspring

#### Gut microbiota of infants

Here, 38 infants born to mothers with SARS-CoV-2 infection during pregnancy or 55 born to healthy controls were analyzed (**Table S3**). We obtained the infant’s stool samples 1-2 days post-partum. Contrary to expectations, SARS-CoV-2 infection during pregnancy did not associate with alpha or beta diversity of the offspring gut microbiota (**Table S4, S5**, and **S6**). However, pre-pregnancy BMI was negatively associated with alpha diversity (**Table S4** and **S5**, Shannon index, Linear regression, P=0.032) and explained most of the microbial composition of the infant’s gut (PERMANOVA, Bray-Curtis dissimilarity, R^2^=4.0%, P=0.025) together with infant weight at birth (PERMANOVA, Bray-Curtis dissimilarity and Sørensen index, R^2^= 3.9%, P<0.020). As expected, the gut microbiota composition varied by delivery mode (PERMANOVA, Bray-Curtis dissimilarity, R^2^=4.7%, P=0.010).

Although the overall gut microbiota composition was unaffected by infection, RFC identified determinant bacterial taxa to be predictive of infants born to pregnant women with SARS-CoV-2 infection. RFC was performed including delivery mode and antibiotic as variables to be selected in addition to taxa. RFC and LIME algorithms showed that gut microbiota of infants from pregnant women with SARS-CoV-2 infections were predicted (F1-score: 0.98) by a higher Lactobacillales (*Enterococcus*) and lower abundance of Pasteurellales (*Haemophilus parainfluenzae*) (**Figure S1B**).

Furthermore, we measured levels of fecal calprotectin in the infant’s stool (meconium). Fecal calprotectin is a non-invasive biomarker that robustly correlates with gut inflammation (*41-46*). Moreover, fecal calprotectin is elevated in infants born to pregnant women suffering from chronic inflammation and is correlated with specific members of the microbiota (*47*). We observed that infants born to pregnant women with active SARS-CoV-2 infection at the time of delivery exhibited a marginally higher fecal calprotectin levels compared to those born to pregnant women with no evidence of SARS-CoV-2 infection or earlier infection (**Figure S3A**, Kruskal-Wallis, P=0.052). We also explored associations of fecal calprotectin levels and taxa abundance. We found that several taxa in active SARS-CoV-2 infections were positively associated with fecal calprotectin. Although only the genus *Leptothrix*, in the healthy control group, was significant (**Figure S3B**, Spearman correlation test, Padj=0.007).

### Oral microbiota of infants

A total of 47 oral samples were obtained from infants: 29 from infants born to SARS-CoV-2 infected pregnant women and 18 born to healthy controls, after 1-2 days post-partum. No alpha diversity significance was observed in infants by their mother’s SARS-CoV-2 diagnosis (**Table S4, S5**, Shannon index and chao1 estimator, Linear regression, P>0.050).

However, infants born to pregnant women with SARS-CoV-2 infection status exhibited a significantly different bacterial composition compared to infants born to healthy controls (PERMANOVA, R^2^=7.9%, P=0.013). Particularly, infants born to pregnant women with active SARS-CoV-2 infection clustered the furthest from those born to healthy controls (**Table S6**, pairwise PERMANOVA, R^2^=11.6%, Padj=0.002). As expected, although to a lesser degree, the delivery mode also affected the oral microbiota composition (PERMANOVA, R^2^=4.6%, P=0.010). We then stratified infants by mode of delivery to eliminate its effect on the microbiota. Here, we observed that infants born vaginally to mothers with SARS-CoV-2 infection exhibited a significantly different microbial composition than those born to healthy controls (**Figure 4A. Table S6**, Bray-Curtis dissimilarity, PERMANOVA, P<0.015). Furthermore, infants born vaginally to pregnant women with early infection presented an oral microbiota that separated the furthest from those infants born vaginally to healthy controls (**Figure 4B, Table S6**, Bray-Curtis dissimilarity, PERMANOVA, P<0.015). No variable significantly explained composition in cesarean-born infants. Interestingly, oral infant microbial composition differed only when assessed by Bray-Curtis dissimilarity, indicating the differences observed were due to changes in highly abundant taxa (**Table S3**).

**Figure 4.**
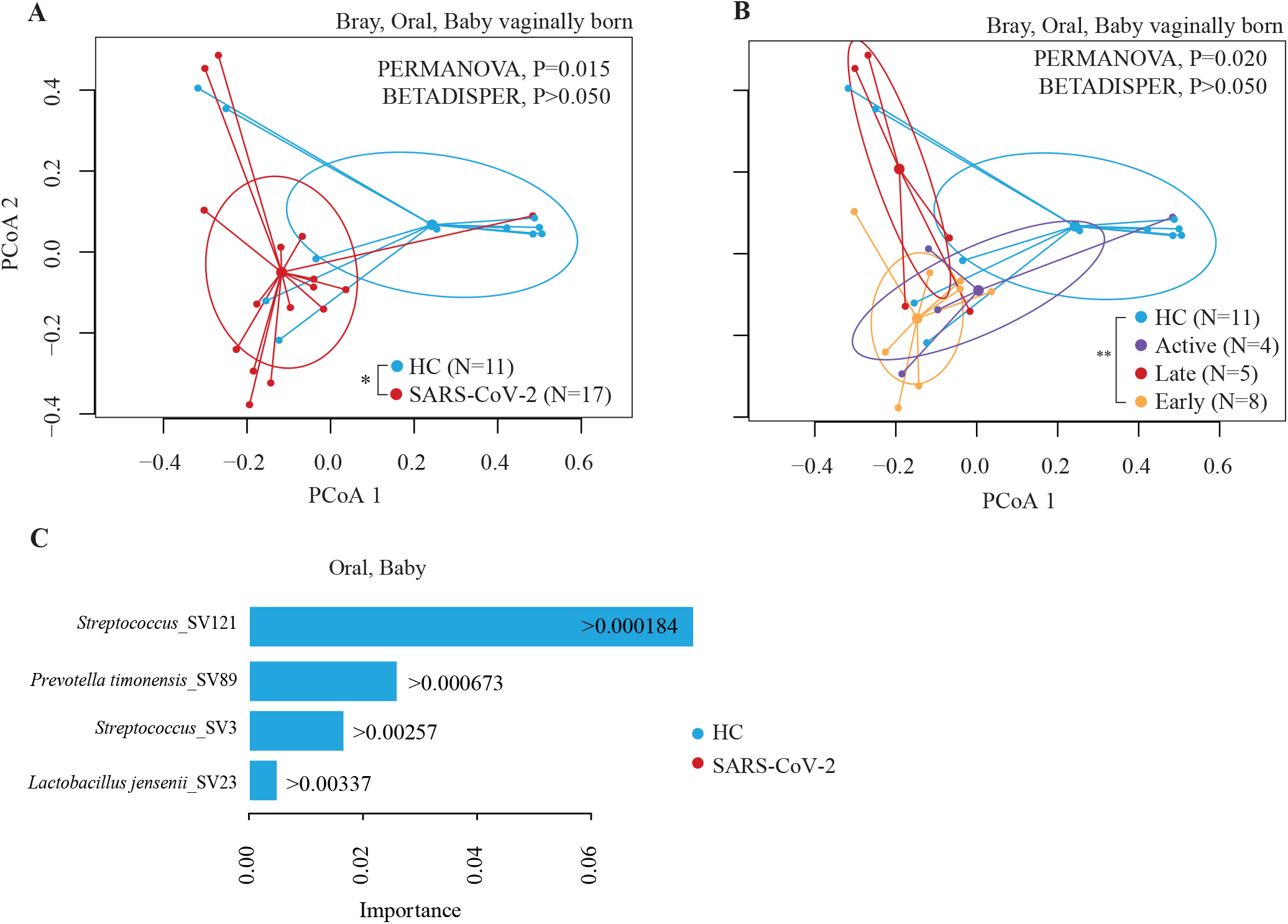
Oral microbial of infants born to pregnant women with SARS-CoV-2 is altered. Beta diversity analyses by groups: (**A**) Infants born to pregnant women infected with SARS-CoV-2 during pregnancy compared to infants born to pregnant healthy controls (HC); (**B**) Infants born to pregnant women infected with SARS-CoV-2 early or late during pregnancy, or with active infection compared to HC. Beta diversity comparisons were performed using PERMANOVA analysis with pairwise comparisons, and BETADISPER for dispersion analysis. For PERMANOVA and BETADISPER analyses we used Sørensen dissimilarities. P values were all adjusted by False Discovery Rate. (**C**) Bacterial taxa (at the amplicon sequence variant or SV) selected by the Random Forest Classification (RFC) and ranked according to their importance in the classification for infants born to pregnant women with SARS-CoV-2 compared to HC. Bars’ colors indicate the comparison group (i.e., SARS-CoV-2 or HC); and each bar indicates the importance by which the increase on an SV predicts a particular comparison group. The selection of the variables for RFC was performed with Boruta algorithm. We also used the Local Interpretable Model-agnostic Explanation (LIME) to estimate a threshold of the abundance of the SV selected with Boruta that predicts a particular comparison group. *Padj<0.050, **Padj<0.010

RFC and LIME algorithms show that infants born to mothers with SARS-CoV-2 infection during pregnancy were predicted (F1-score: 0.99) by higher abundance of Bacteroidales (*Prevotella timonensis*) and low abundance of Lactobacillales (*Streptococcus* and *Lactobacillus jensenii*, **Figure 4C**).

### Pregnant women with early and late but not active SARS-CoV-2 infections transferred viral antibodies to their infants

Finally, we measured IgG antibody levels against nucleocapsid protein (NP) in cord blood to determine whether maternal antibodies were vertically transferred to their infants. We found significantly higher IgG levels in cord blood from pregnant women with early and late infections but not with active infection compared to healthy controls (**Figure S2**).

## DISCUSSION

We observed that SARS-CoV-2 infection during pregnancy, particularly in early pregnancy or active infection at time of delivery, resulted in perturbations of the vaginal, gut, and oral microbiota of pregnant women. Moreover, the microbiome alterations in pregnant women were reflected in the infant’s oral microbiome. To our knowledge, this is the first study exploring the impact of SARS-CoV-2 infection on the vaginal and oral microbiota of pregnant women and their effect on the offspring’s gut and oral microbiota.

First, we observed a decrease in gut microbial diversity in infected pregnant women. Low gut microbial diversity has been linked to negative health outcomes (*48, 49*). The fact that these changes were observed even if the infection was early in pregnancy, suggests the long-lasting effect of SARS-CoV-2 infection on gut microbial diversity (*38*). Similarly, other studies on non-pregnant Chinese population, COVID-19 is also associated with reduction in gut microbial richness (*33*) even lower than in H1N1 hospitalized patients (*35*), and also reported a long-lasting effect for at least 6 months post-infection (*33*). Yet, a recent study including pregnant women from Mexico whose stool samples tested positive for SARS-CoV-2, despite of being asymptomatic or negative on nasopharyngeal swabs, suggesting earlier infection, do not report statistical differences in gut microbiota alpha diversity but showed differences in the gut microbiota composition and in specific taxa (*38*). Differences in results, might be due to the presence of SARS-CoV-2 in the gut - we did not assess SARS-CoV-2 in stools; or to geographical differences in the microbiota that could result in a varying response to the viral infection. Moreover, we also observed that infants born to women with active infection presented elevated fecal calprotectin levels indicative of gut inflammation. In our study, only *Leptothrix* (a non-pathogenic naturally occurring gut genus) was positively associated with fecal calprotectin in healthy controls, however, no association was observed for any of the infants born to mothers with SARS-CoV-2 during pregnancy. Differences in the immunological states of babies born to mothers with past SARS-CoV-2 infections compared to healthy controls may be shaping this correlation.

A recent study from Spain reported that the overall composition of the nasopharyngeal microbiota differs in pregnant women with SARS-CoV-2 infection compared to healthy controls (*37*). Specifically, the authors observed a higher abundance of Prevotellaceae family (Bacteroidales order) in pregnant women with active SARS-CoV-2 infection. The SARS-CoV-2-infected pregnant cohort included in this study, showed prediction by the higher abundance of members of the Prevotellaceae family, in the oral cavity. Members of this bacteria family such as *P. intermedia*, are considered the main bacterial specie implicated in acute periodontal lesions. Similarly, *H. parainfluenzae* an oral commensal associated with beneficial immunomodulatory effects is decreased in the pregnant women with SARS-CoV-2 included in this study (*50*). Conversely, *F. nucleatum*, decreased in pregnant women with SARS-CoV-2 infection in our study, is one of the most prevalent species and by far the most prevalent oral species implicated in adverse pregnancy outcomes (*51, 52*). Overall, our results suggest that SARS-CoV-2 infection plays an important role in dictating the abundance of bacteria linked to immune regulation and pregnancy outcomes.

In terms of the vaginal microbiota, different studies have reported a stable vaginal microbiota composition even during pregnancy, particularly in the Caucasian population (*53, 54*), which is most of the participants of this study. We demonstrate that SARS-CoV-2 infection impacts the vaginal microbiota richness and composition. Compositional vaginal microbiota changes observed in pregnant women with active SARS-CoV-2 infection may be the consequence of alterations in the vaginal epithelial environment (*55*) and the immune system interaction. Although viral particles of SARS-CoV-2 have not been detected in the vaginal fluid (*56-58*), this pulmonary infection promotes strong systemic inflammatory responses (*59*). This pro-inflammatory immune tone on the epitheliums, including the vagina, may limit or favor the survival of certain taxa. Furthermore, we also observed a higher vaginal microbial heterogeneity among infected women compared to controls. This high heterogeneity was previously observed in a metagenomic study with post-menopausal women (*56*), where not only microbial but also the proportion of bacterial transcript varied considerably among SARS-CoV-2 infected participants in that study (*56*). Such inter-individual variation may be due to the differences in the personal physiological response to SARS-CoV-2, disease severity, individual’s hormonal profile (e.g., early, or late in pregnancy), and health status. Vaginal low microbial richness is frequently associated with a healthy state opposite to the high richness found in bacterial vaginosis. Contrary to expected, mothers with SARS-CoV-2 infection exhibited low vaginal microbial richness compared to healthy controls, with a decrease of several bacterial taxa associated with bacterial vaginosis. The clinical implications of this decreased richness in the vaginal microbiota of SARS-CoV-2 pregnant women deserve further investigation.

Furthermore, the effect of SARS-CoV-2 infections on the pregnant women’s microbiota was further reflected in the offspring born vaginally. The mother’s vaginal and rectal microbiota is the major colonizer of the infant’s gut and oral microbiome (*60-63*). Accordingly, bacterial species expected to be the first colonizers of the infant’s oral cavity (*64*), were among the ones affected by SARS-CoV-2 infection in vaginally delivered infants. The composition of the oral microbiome is established early in life, is stable throughout life (*65, 66*), and it has implications for long-term health (*67, 68*). Despite oral microbiota changes in infants, we did not observe an effect of SARS-CoV-2 infection during pregnancy on the offspring’s gut microbiota, similar to previously reported (*69*). However, we observed that infants born to pregnant women with active infection presented elevated fecal calprotectin levels indicative of a pro-inflammatory tone early in life. Elevated fecal calprotectin has been also seen in infants born to pregnant women with inflammatory bowel disease (IBD). IBD is characterized by dysbiotic microbiota. In line with this, gut abundance of *Faecalibacterium, Bifidobacterium*, and *Alistipes* have shown to have negative correlations with levels of fecal calprotectin within 3 months of birth while *Streptococcus* is positively correlated with fecal calprotectin levels.

Finally, we found that pregnant women with active SARS-CoV-2 infection were more likely to deliver by Cesarean section. We hypothesize increased rates of Cesarean section in this cohort, might be due to the deteriorating health of a pregnant woman with active SARS-CoV-2 infection and/or the inability of an infant to tolerate labor with illness.

In the clinic, it is important to recognize the effect of SARS-CoV-2 infections in mothers and its influence on infants. By simply knowing there are changes that occur with SARS-CoV-2 infection and inflammation caused by this infection, we can begin to understand the implications beyond immediate infection risks. As alterations in the microbiota can have health implications in the infant, these changes are important to characterize, and potential treatment can be explored to counter these changes. For example, besides avoiding SARS-CoV-2 infections, actions such as probiotic intake and a diet focused on microbiota balance might be a benefit for the already infected patients. Main attention might be given to the parents since they will be the principal source of microbiota colonization of the infant. As COVID-19 becomes more endemic and as the COVID-19 vaccine is widely recommended in pregnancy, it will be important to address all risks to SARS-CoV-2 infection, which with future research may include changes in the gut microbiota and possible therapies to prevent such changes.

## LIMITATIONS

Study limitations include low sample size, particularly for infants, which stratification by vaginally born and cesarean greatly reduced the power to discriminate between SARS-CoV-2 infection and healthy control groups. Additionally, participants were mostly Caucasian, followed by Hispanic, which limits the conclusions to this population sector, particularly important for vaginal microbiota which is highly dependent on the racio-ethnic background (*40*). Additionally, the use of antibiotics during delivery was present in an important part of the population (37%), mostly for the standard treatment of group B streptococcus when present, and although it was similarly distributed among groups, the effect on non-antibiotic users would be important to investigate.

We also had limited diversity of controls, as some controls were recruited pre-pandemic for another study. Therefore, reporting on both demographics and perinatal outcomes has limited relevance in this manuscript, as specifically controls were chosen out of convenience of the timing of delivery and sample collection, often a scheduled cesarean delivery, unlike cases who were approached when identified as having had prior SARS-CoV-2 infection and presenting to the labor floor. Ultimately, despite these limitations, our study conveys differences in the microbiota of the birthing parent, which will need to be explored in future research studies.

Limitations in the frequency and availability of SARS-CoV2 diagnostics during the first year of the COVID-19 pandemic when our study was conducted may have led to underreporting infections during pregnancy. However, our antibody results, using cord blood as a proxy for the presence of maternal antibodies to NP IgG aided proper classification of our study participants.

## MATERIALS AND METHODS

### Patient recruitment

We enrolled pregnant women and their newborns delivered at the University of Massachusetts Memorial Hospital between April 2020 and August 2021. A total of 100 pregnant women, 62 with SARS-CoV-2 diagnosis, 38 without SARS-CoV-2 diagnosis, and 77 newborns (2 sets of twins) were recruited (**Table 1**). Ten of these control cases were recruited prior to the pandemic as part of an ongoing MELODY trial (*70*) (docket # H00016462#). Participants were classified as having had SARS-CoV-2 infection by clinical PCR positive viral DNA diagnostic test at any time during pregnancy or as healthy controls if no positive diagnostic test was either listed in their medical record or reported by the patients, and they tested negative upon admission to labor and delivery, as per hospital protocol. We also, defined negatives to SARS-CoV-2 by ELISA, as previously defined by us (*39*). Briefly, a maximum specificity threshold was established based on a cutoff at 100% specificity for the NP IgG assessed on pre-pandemic samples collected from 96 adult healthy individuals. The cutoff is an OD value of 0.57 and represents the highest OD value for NP IgG in those pre-pandemic adult healthy individuals (*39*).

SARS-CoV-2 positive participants were further sub-classified by the time of SARS-CoV-2 diagnosis in “early” (1^st^ – 2^nd^ trimester), “late” (3^rd^ trimester), or “active” (SARS-CoV-2 positive at delivery) groups. Mother-infant dyads were incomplete when samples could not be taken or processed, or the parents only consented to the collection of either mother or infant samples. This cohort was recruited under the COVID-19 Analysis on Perinatal Specimens Related to Exposure (CARES) protocol (docket # H00020145), with the additional10 healthy pregnant women and their newborns recruited as part of our ongoing MELODY trial (*70*). The Institutional Review Board at the University of Massachusetts Chan Medical School approved both studies. Informed consent was obtained from all study participants or their health care proxy using REDCap digital signatures to reduce the potential for patient-staff transmission of SARS-CoV-2 or based on the remote data collection study design for the MELODY project. Participants were asked to consent separately to each of these sample collections, and therefore not all participants had samples in each cohort.

### Sample collection

After obtaining consent, all samples were collected by the attending or resident physician or nurse caring for the patient at delivery time. From pregnant women, we obtained anal, oral, and vaginal swabs before delivery. For anal samples, a sterile swab (Sterile Flock Swab. Puritan Medical Products Company LLC, ME, US) was inserted 1 to 2 inches into the anus to obtain gut material from pregnant women prior to delivery. Oral swabs were obtained using the OMNIgene•ORAL (DNAGenotek™, Canada) following the manufacturer instructions. Briefly, the oral mucosa was sampled from the tongue for 30 seconds. We used the OMNIgene•VAGINAL (DNAGenotek™, Canada) to obtain vaginal samples prior to delivery. Specifically, a sterile swab was inserted 1 to 2 inches into the vagina and rotated in circles along the vaginal walls for 20 seconds. After swabbing, both oral and vaginal swabs were inserted into their respective tubes containing a DNA/RNA stabilizer buffer. For antibody assays, we collected cord blood after delivery, namely: 5 cc of blood were withdrawn or drained into EDTA tube. Plasma was separated from peripheral blood cell pellet by centrifugation, 10 minutes, room temperature and aliquots stored at -20 ºC until thawed for antibody testing. For the newborns, we collected samples at 1-2 days after delivery; namely, a diaper with the meconium as previously described and an oral swab as described above.

### Clinical data

All clinical data were obtained retrospectively by reviewing the electronic medical records of each participant following delivery.

### Nucleic acid isolation

To minimize the risk of SARS-CoV-2 infection, oral samples were inactivated at 65-70 ºC for one hour, as demonstrated elsewhere (*71*). Then, samples were pre-treated with Proteinase K (Cat # P8107S, New England Biolabs, MA, USA) and incubated for 2 hours at 50 ºC, and subsequently used for nucleic acid isolation. Nucleic acid isolation of oral samples and mother’s anal swabs were performed using the ZymoBIOMICS DNA/RNA Miniprep Kit (Cat # D7003/D7003T, Zymo Research, CA, USA) following the manufacturer recommendations for parallel isolation of DNA and RNA. Nucleic acid from the meconium of the newborns was isolated using DNeasy Power Soil Pro kit (Cat # 47016, Qiagen, CA). Due to the tar-like consistency of the meconium, a combination of bead-beating, 30 minutes of heating at 80ºC, and only 90-100 mg of sample was used for the initial lysis step.

### Microbiome profiling

The *16S rRNA* gene was sequenced following methods previously described (*71*) using the 341F and 806R universal primers to amplify the V3-V4 region. The 300nt paired-end sequences were generated on the Illumina MiSeq platform. Replicate reactions were performed for each sample and the read data were merged for analysis. Only, forward *16S rRNA* gene MiSeq-generated amplicon sequencing reads were dereplicated and sequences were inferred using DADA2 (*71*). We obtained in average 57,428 (±43,107) sequences per sample. Generated sequences were deposited in the NCBI database, BioProject ID: PRJNA871082. Potentially chimeric sequences were removed using consensus-based methods. Taxonomic assignments were made using BLASTN against the NCBI refseq RNA database combined with GreenGenes, the Human Oral Microbiome Database, and cervicovaginal microbiome *16S rRNA* reference sequences from NCBI previously used (*72*). These files were imported into R and merged with a metadata file into a single Phyloseq object. Samples were rarefied at 4,085 sequences per sample (**Table S8**).

### Community State Types (CSTs)

Each women’s sample was classified into CST following the protocol of the VALENCIA program (*73*) with Python 3 (https://github.com/ravel-lab/VALENCIA). Input data was formatted using local scripts.

### Random forest classification (RFC)

RFC was used to find microbiome and clinical variables that could predict SARS-CoV-2 infection. First, the feature selection was run, in which the wrapper Boruta (*74*) is used to identify a subset of covariates that is predictive of the outcome, followed by RFC utilizing only the Boruta-selected subset. For the RFC interpretation, the models were entered into the Local Interpretable Model-agnostic Explanation (LIME) toolbox (*75*). LIME allows identified human-interpretable logical rules on the microbiome to distinguish between patients with different outcomes.

### Antibody ELISA

Antibodies against the receptor binding domain (RBD) of the SARS-CoV-2 spike protein were measured by ELISA following published methods (*76*). In brief, an IgG antibody against the nucleocapsid protein (NP, gifts from Lisa Cavacini, UMass-Biologics) was used at 0.5 µg/mL and incubated with plasma at a 1:100 dilution. Optical density (OD) was measured at 450 nm and 570 nm on the SpectraMax iD5 ELISA plate reader (Molecular Devices) using SoftMax Pro software (version 7.1, Molecular Devices). For the positive antibody control, monoclonal therapeutic CR3022 IgG antibody (gifts from Lisa Cavacini, UMass-Biologics) was diluted from a concentration of 2.5 μg/ml in dilution buffer to 12 two-fold serial dilutions to generate the standard control curve (*77*). The 570 nm OD was subtracted from the 450 nm OD for the final OD value. Antibody levels were used as a continuous variable in the analysis.

### Statistical analysis

Fisher’s exact test, T-test or Kruskal-Wallis were used to evaluate differences in demographics among pregnant women with different SARS-CoV-2 statuses by pregnancy stage (SARS-CoV-2 positive/HC, and Early/Late/active SARS-CoV-2 infection/HC). For the microbiota diversity analyses, comparisons were performed between groups at ASV level using multiple functions from Phyloseq v1.19.1 package (*78*) in R (*79*). Microbial alpha diversities were measured using Shannon index or Chao1 estimator. Linear regressions (LM) were run with “lm” base R function. Alpha diversity metrics were set as the dependent variables, while SARS-CoV-2 infection (and its different stratifications), race, antibiotic use, mother’s age, pre-pregnancy body mass index (BMI), gestational diabetes, and delivery mode (the last one only for infant samples) as the independent variables. Best-fitted models were chosen using the “step” R function. The “emmans” function (*80*) was used as a pairwise analysis since it computes contrasts, trends, and comparisons of slopes among groups. Beta diversity was evaluated using Bray-Curtis dissimilarity and Sørensen index; the first one considers the ASV abundance while the second only the incidences. The same variables as for alpha diversity analysis were included in this model. Groups were analyzed using non-parametric Permutational Multivariate Analysis of Variance (PERMANOVA) (*81*) with “adonis2” from vegan package (*82*) and pairwise analyses were performed with “pairwiseAdonis” (*83*). PERMANOVA allows comparing variance between groups to the variance within groups (spatial location differences). Sample dispersion was also evaluated using PERMDISP2 procedure (*84*) with “betadisper” function, which executed the analysis of multivariate homogeneity of group dispersions (variances), adjusting for the different sample sizes to avoid bias. Association between microbial and fecal calprotectin values was assessed by the spearman correlation test with “cor.test” function on base R. The correlation analysis excluded genera with a mean abundance of <0.1% or more than 80% zero values as previously performed by other authors (*47*). P values were all adjusted with the false discovery rate method using the “p.adjust” function from R base. Plots were generated using the “ggplot2” (*85*) package and base R functions and edited in Adobe Illustrator (*86*).

## Data Availability

Sequencing data are available in a public, open-access repository: the NCBI database, BioProject ID: PRJNA871082. Additional de-identified clinical metadata are available on reasonable request.

https://dataview.ncbi.nlm.nih.gov/object/PRJNA871082?reviewer=9jjclf6enqcubg06bf88d65pmb

## Acknowledgments

We thank all the pregnant women that agreed to participate in the study and provided their samples, their infant samples, and time to complete the study. We also acknowledge the help of nurses on the delivery floor and Devon Holler in the lab, both aiding sample collection and transportation from the hospital to the laboratory. We also thank Rafael López Martinez for his support in data handling and processing.

## Author contributions

Conceptualization: AMC and HKL

Methodology: HKL, AMC, MRC, DVW, AMM, GF, CF, VB, SB, YYR

Investigation: HKL, AMC, MRC, DVW, AMM, GF, CF, TY, AC, AK, AA, and DVR

Visualization: DVR, VB

Funding acquisition: AMC, HKL, and AMM

Supervision: AMC and HKL

Writing – original draft: HKL, DVR, AMC

Writing – review & editing: all authors

## Data availability statement

Sequencing data are available in a public, open-access repository. Additional de-identified clinical metadata are available on reasonable request.

## SUPPLEMENTARY FIGURES

**Figure S1.**
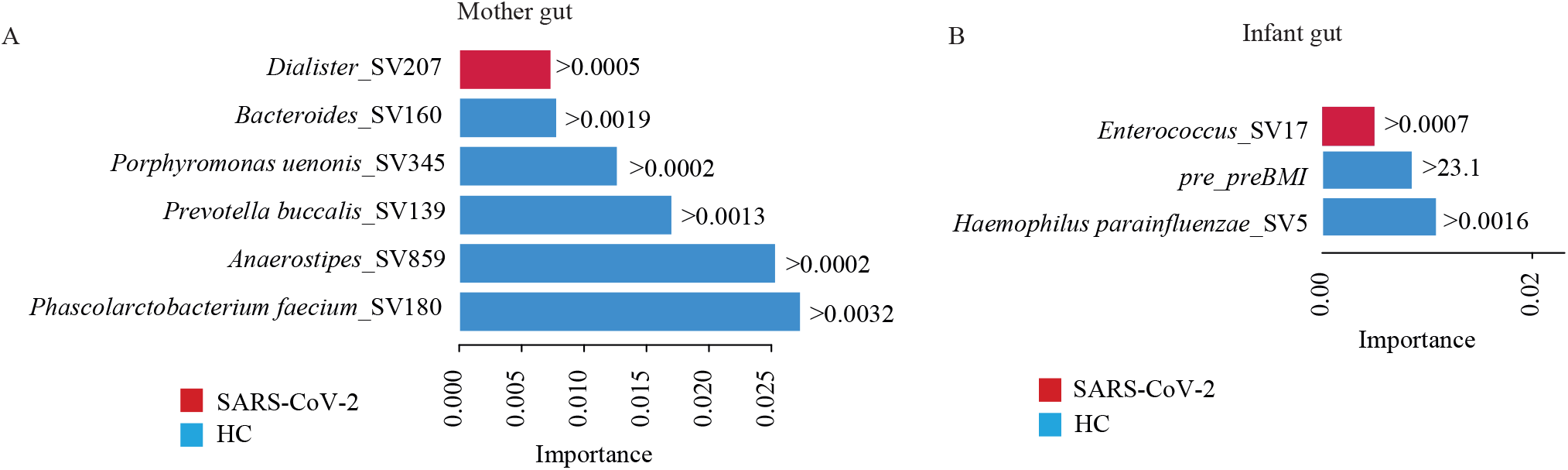
Bacterial taxa (at the amplicon sequence variant or SV) for mother and infant gut samples selected by the Random Forest Classification (RFC) and ranked according to their importance in the classification. RFC is shown in: (**A**) Pregnant women with SARS-CoV-2 infection during pregnancy vs. HC, (**B**) Infants born to mothers with early SARS-CoV-2 infection during their pregnancy vs. HC. Each bar indicates the importance of a particular SV predicting a comparison group. The selection of these variables for RFC was performed with Boruta algorithm. Values inside or next to the bars correspond to the abundance threshold above which a particular SV predicts for a comparison group, shown in colors (i.e., red for SARS-CoV-2 or blue for HC). We used the Local Interpretable Model-agnostic Explanation (LIME) to estimate these abundance thresholds and therefore the direction of the SASR-CoV-2–microbiota associations.

**Figure S2.**
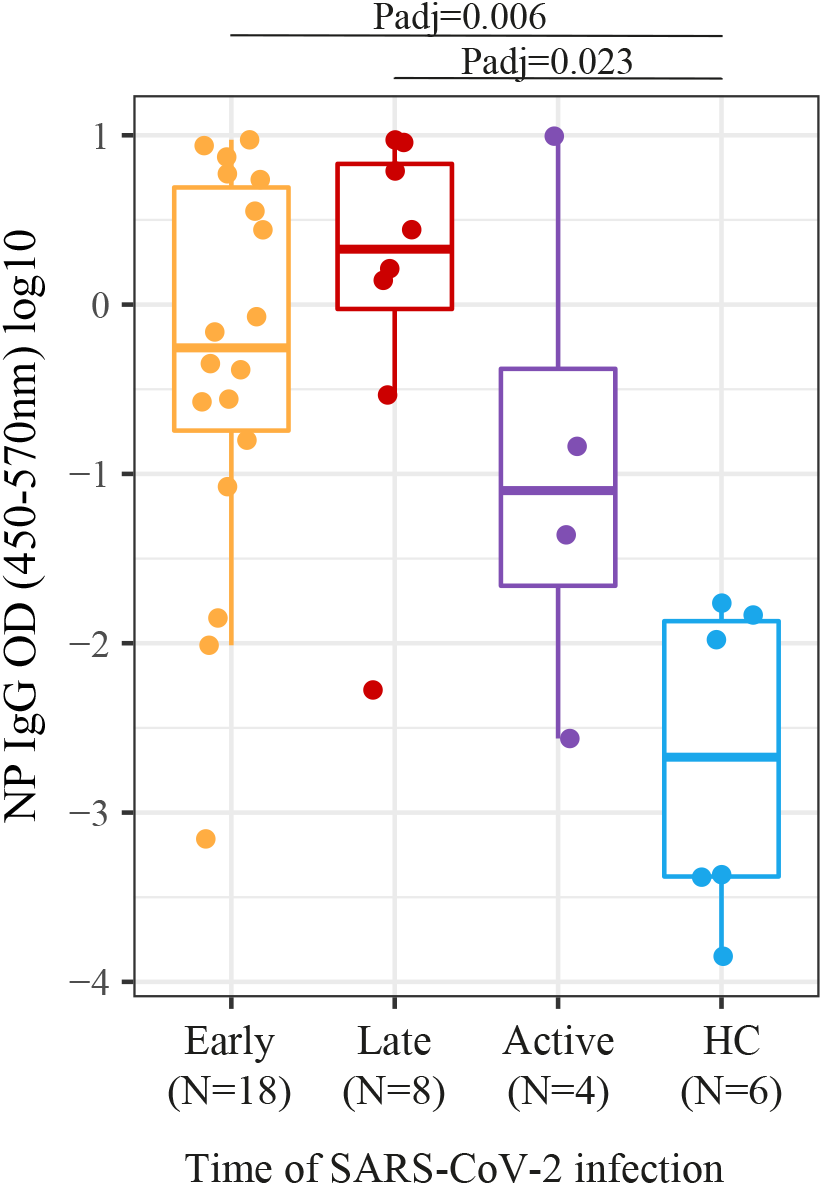
IgG antibody levels against nucleocapsid protein (NP) in cord blood in women with SARS-CoV-2 infection during pregnancy and healthy controls (HC).

**Figure S3.**
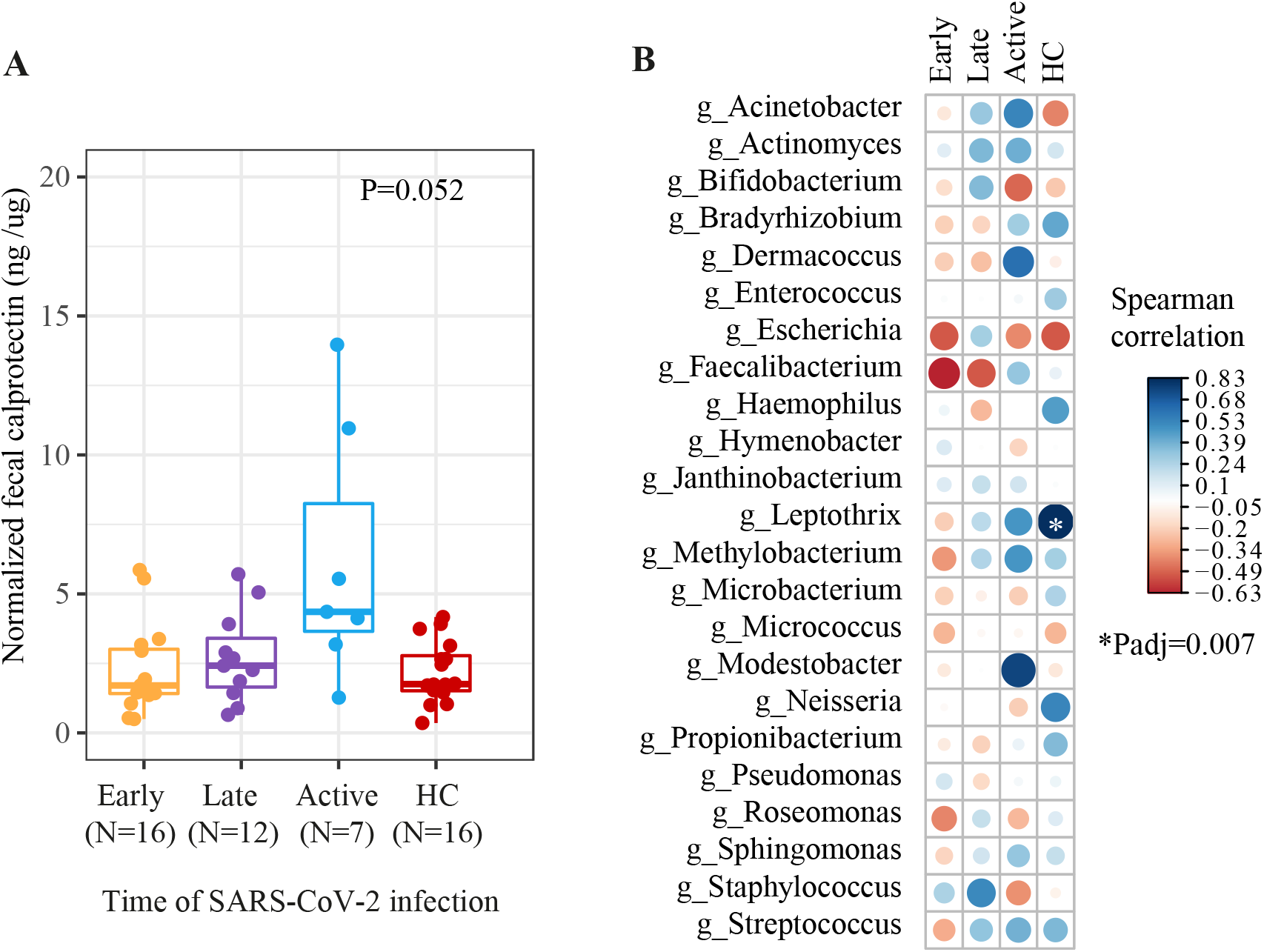
Normalized fecal calprotecting levels for infant’s gut samples. (**A)** Normalized fecal calprotecting levels among infants born to pregnant women with SARS-CoV-2 infection at different times during pregnancy compared to healthy controls (HC). Kruskal-Wallis test was performed. (**B**) Spearman correlation between infant fecal calprotectin and taxa abundance at the genus level. Significance was only reached by *Leptothrix* genus in the HC group. Blue and red colors indicate positive and negative correlations respectively. Size of the circles indicate the level of a particular correlation: big circle, high correlation; and small circle, low correlation. P value adjustment for multiple comparison was performed with the “false discovery rate” method.

